# Epidemiological Interactions of Influenza and SARS-CoV-2 within a University Population During Omicron B.1.1.529 Outbreak

**DOI:** 10.1101/2022.05.26.22275641

**Authors:** Rachel Christine Malampy, Troy James Ganz, Gabriel M DeOliveira, Catherine T Le, Kurvin M Li, Jared Auclair

## Abstract

The COVID-19 Pandemic has prompted innovation and research to further understand not only SARS-CoV-2, but other respiratory viruses as well. Since the start of the pandemic there has been a lack in influenza collection and surveillance. In October 2021 the Life Sciences Testing Center at Northeastern University implemented the TaqPath™ COVID-19, Flu A, Flu B combo kit to test for multiple respiratory diseases among the University’s population. During this time the SARS-CoV-2 variant of concern, Omicron B.1.1.529, became the dominant strain in the greater Boston area. During this time an inverse correlation in the detection of positive SARS-CoV-2 and Influenza A was observed. More data is needed to determine if this observed inverse correlation on positivity rate is linked to public health measures or biological mechanism within the immune system.

## Introduction

Infectious disease epidemiology has recently suffered operational burdens as the coronavirus disease (COVID-19) pandemic has inexorably diverted resources from routine monitoring [1]. Continuous surveillance of influenza virus has been vital for vaccine production. As enveloped RNA viruses that target, and transmit via, the respiratory system, severe acute respiratory syndrome coronavirus-2 (SARS-CoV-2) and influenza infection present in similar symptoms of fever, headache, pharyngitis and tracheobronchitis. They distinguish themselves on a mechanistic basis as SARS-CoV-2 interacts with angiotensin converting enzyme 2 (ACE2) receptor on epithelial and endothelial tissue for cell entry, while influenza associates with sialic acid groups on surface glycoproteins of epithelial cells [2].

## Results

From November 1^st^, 2021, to February 28^th^, 2022, 3065 individuals were tested, with the first influenza A detected on November 4, 2021, reaching an average weekly prevalence rate of 13.62%, 95% CI [10.88,16.35] (Fig. 1A). A sharp decrease of Influenza A cases at a rate of 12.5% was observed between December 27, 2021, and January 7, 2022, while SARS-CoV-2 cases increased at a rate of 3.75% during the same period (Fig. 1B-C). With news of the rapidly spreading Omicron (B.1.1.529) variant, samples reactive for SARS-CoV-2 were crossed referenced on the TaqPath™ COVID-19 Combo Kit, whose RT-qPCR targets permit proxy identification via S-gene target failure (SGTF) [3]. Retrospective analysis indicated a demonstrable suppression of influenza cases with the surge in Omicron B.1.1.529 (Fig. 1D).

**Figure 1:**
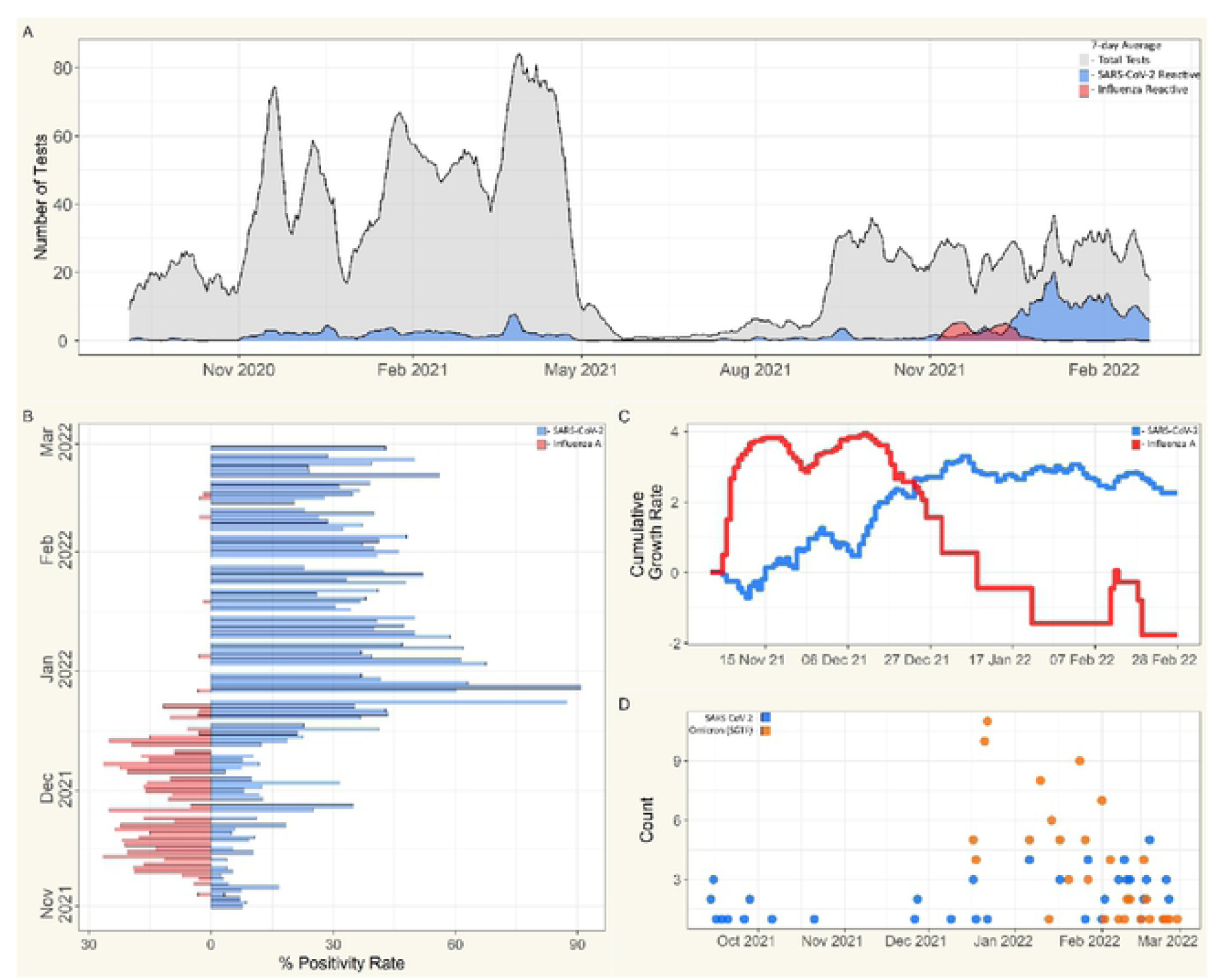
A) Seven-day averages for RT-qPCR testing collected from symptomatic individuals; tests collected, **grey**, reactive for SARS-CoV-2, **blue**, reactive for Flu **A, red. B)** Positivity rate for symptomatic cohort from November 1^st^, 2021,. to February 28^th^, 2022; SARS-CoV-2, **blue**; Flu A. **red**. C) Cumulative growth rates for seven-day averages of SARS-CoV-2 and influenza incidence in symptomatic cohort from November 1^st^. 2021. to February 28^th^, 2022; SARS-CoV-2. **blue**; Flu A. r**ed**. D) Totals of SARS-CoV-2 Omicron (B.1.1.529) variant determination via S-gcnc target failure (SGTF) for symptomatic population from September 14^th^, 2021, to February 28^th^ 2022; SARS-CoV-2, **blue**; SGTF, **orange**, n=172.

## Discussion

Our observation supports possible SARS-CoV-2 dominance via unique host interactions as well as the potential occurrence of priming the immune system to limit co-infection [4]. Symptomatic testing demand was higher during the first year of the pandemic with weekly averages of 38.5, 95% CI [35.8, 41.2], daily orders, yet the weekly average SARS-CoV-2 positive cases of 1.6, 95%CI [1.4, 1.7] may further indicate the activity of another pathogen during that time. To date, we have reported a single co-infection of COVID-19 (Ct:19.8) with Flu B (Ct:38.9) and admit a scarcity in the literature regarding observations of viral interactions in human populations.

Selective pressures derived from competitive relationships may impact virulence or seasonality and, during phases of impeded surveillance, other factors such as zoonotic transmission and Long-COVID may warrant consideration [5,6]. The inverse relationship SARS-CoV-2 and influenza demonstrated in this analysis advocates for health care providers to employ more complete respiratory panels for suspected viral infections to aid in monitoring of strain dominance and co-infection.

## Materials and Methods

Our lab, the Life Sciences Testing Center (Burlington, MA), has performed SARS-CoV-2 molecular diagnostics for the Northeastern University population since August 2020 using the TaqPath™ COVID-19 Combo kit (Thermo Fisher Scientific, CAT#: A47814). As previously reported, collection of anterior nare swabs was conducted at facilities designated for either symptomatic or non-symptomatic individuals [3]. Individuals were directed to submit a sample at the symptomatic site based on whether they were experiencing COVID-19-related symptoms or were close contacts of a COVID-19 case. On October 25^th^, 2021, we implemented the TaqPath™ COVID-19, FluA, FluB Combo Kit (Thermo Fisher Scientific, Cat#: A49868), a multiplex RT-qPCR assay, to monitor COVID-19, Influenza A, and Influenza B among the symptomatic cohort. Individuals who tested positive for SARS-CoV-2 were required not to test for 90 days post infection, which prevented data collection bias.

## Data Availability

All relevant data are within the manuscript and its Supporting Information files.

## Notes

### Competing Interest Statement

The authors have declared no competing interest.

### Funding Statement

The author(s) received no specific funding for this work.

### Author Declarations

Northeastern University IRB

